# Exploring the utility of social-ecological and entomological risk factors for dengue infection as surveillance indicators in the dengue hyper-endemic city of Machala, Ecuador

**DOI:** 10.1101/2020.08.31.20185355

**Authors:** Catherine A. Lippi, Anna M. Stewart-Ibarra, Timothy P. Endy, Mark Abbott, Cinthya Cueva, Froilán Heras, Mark Polhemus, Efraín Beltrán-Ayala, Sadie J. Ryan

## Abstract

The management of mosquito-borne diseases is a challenge in southern coastal Ecuador, where dengue is hyper-endemic and co-circulates with other arboviral diseases. Prior work in the region has explored social-ecological factors, dengue case data, and entomological indices. In this study, we bring together entomological and epidemiological data to describe links between social-ecological factors associated with risk of dengue transmission at the household level in Machala, Ecuador. Households surveys were conducted from 2014–2016 to assess the presence of adult *Aedes aegypti* (collected via aspiration) and to enumerate housing conditions, demographics, and mosquito prevention behaviors. Household-level dengue infection status was determined by laboratory diagnostics in 2014–2015. Logistic models and multimodel selection were used to identify social-ecological variables associated with household presence of female *Ae. aegypti* and household dengue infection status, respectively. We identified significant risk factors for household-level dengue infection reflecting household condition, including bamboo cane construction material, shaded patios, and adjacency to abandoned properties, while housing structures in good condition were identified as protective against dengue infection. *Aedes aegypti* presence was associated with a greater than average number people per household and interrupted water supply, but was not associated with household level dengue infections. Models of *Ae aegypti* presence were unstable, and not well resolved in ranking of competing models, suggesting that highly localized entomological surveillance indicators may not be indicative of risk in communities with hyperendemic dengue fever. These findings add to our understanding of the systems of mosquito-borne disease transmission in Machala, and in the larger region of southern Ecuador, aiding in the development of improved vector surveillance efforts, and targeted interventions.

## Introduction

The management of arthropod-borne viruses (arboviruses) is a fundamental task of public health agencies throughout much of Latin America and the tropics. Pathogens transmitted by the mosquito *Aedes aegypti*, such as dengue virus, impose some of the greatest growing disease burdens in this region [1]. Dengue has increased 30-fold over the past 50-years, globally and in South America, and over the past decade more than 19.6 million cases of dengue infection were reported in the Americas [2-4]. Dengue, like many arboviral infections, has limited options for clinical treatment and marketable vaccines, leaving vector control as a primary means of preventing and mitigating large outbreaks [5-7]. While there are many established protocols for designing and conducting public health vector control activities, these are subject to logistical constraints, resource availability, and fluctuating funding streams [6-11]. Development of targeted vector control strategies that focus on surveillance indicators or known risk factors for transmission offer a possible solution for implementing effective control measures in resource-limited communities (e.g., Lippi et al. 2020) [12]. Yet, social and ecological risk of exposure to arboviral infections is often understudied at fine spatial scales at which most vector control interventions operate, where household-level features and behaviors can vary greatly within the built environment [13-15], leading to varying risk of exposure to vectors, and thus, transmission.

Entomological and epidemiological surveillance systems are integral components of public health vector control programs, where incoming data are key for informing management strategies and developing policy [11,16,17]. Vector abundance data are often used by public health agencies to trigger vector control responses, though the utility of directly inferring transmission risk from *Ae. aegypti* densities is difficult to establish, and varies widely across surveillance programs [17–19]. Beyond informing public agency responses, surveillance indicators can be used in the broader context of modeling risk by establishing correlative links between health outcomes and the social-ecological conditions in which they occur [20,21]. Ecological studies that establish such connections are routinely seen in the public health literature [22-24]. While observational studies are inappropriate for inferring causality, they remain useful for informing current decision-making and guiding further epidemiological investigations.

In Ecuador, the national public health agency, the Ministerio de Salud Pública (MSP), oversees vector control and surveillance initiatives [25,26]. Ecuador’s southern coastal region is an area of historically high mosquito-borne disease activity [27,28]. El Oro province, located on the border with Peru, is consistently burdened with high case numbers of dengue, where four serotypes of dengue virus are in circulation (DENV1-4), and more recently, chikungunya and Zika viruses [27]. The port city of Machala, with an estimated population of over 280,000, is Ecuador’s fourth largest city, the capital of El Oro province, and a major epicenter of commercial agricultural trade (Fig. 1) [29].

**Figure 1.**
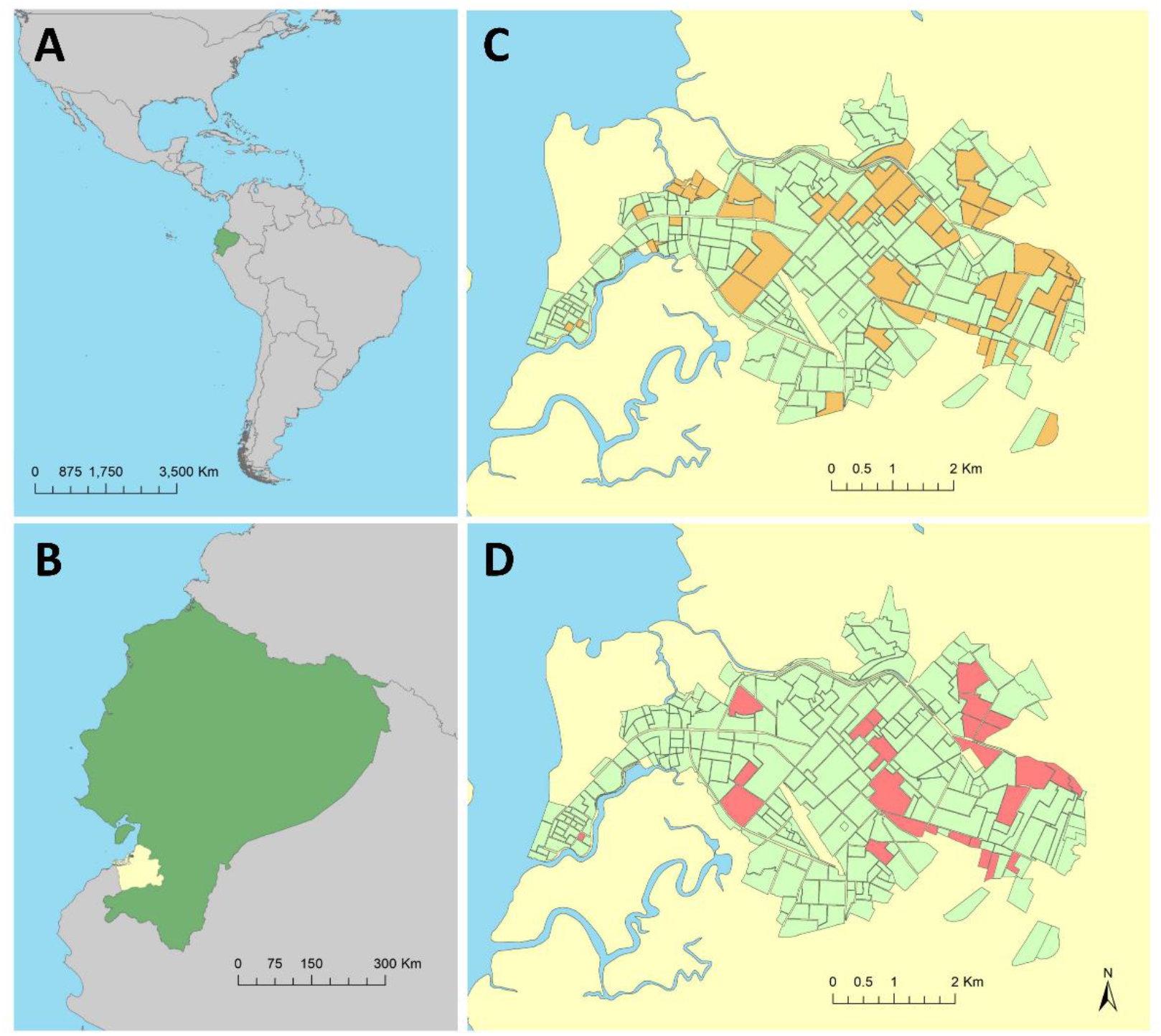
The study was conducted Machala, a city in the South American (A) country of Ecuador (B), located in El Oro province (B, shown in yellow). Households sampled during this study were located throughout the city of Machala, consisting of 443 households in 91 sampling clusters surveyed for *Aedes aegypti* mosquitoes (C), and 141 households in 33 sampling clusters surveyed for both *Ae. aegypti* mosquitoes and dengue infection status (D). Household locations were aggregated to census block for de-identification purposes in these figures.

Dengue is hyperendemic in Machala, where *Ae. aegypti* is prevalent and transmission can occur throughout the year [30,31]. The high incidence of dengue, large population, and longstanding history of vector control and surveillance activities make Machala an ideal setting for studying arboviral transmission systems. Several previous studies conducted in Machala have assessed risk factors for dengue fever outbreaks at the mesoscale (e.g. neighborhoods or census including vector densities, environmental drivers, sociodemographic characteristics, community perceptions, and service accessibility [12,14,25,30,32–34]. The findings of these studies are useful in a public health advisory capacity, providing information that guides decision-making and research. Nevertheless, studies conducted on data aggregated to administrative units face limitations in terms of guiding spatially explicit interventions on fine scales, and we cannot assume that neighborhood-level associations will be observable at the level of households, where people are exposed to mosquitoes [35-37], and at which scale interventions such as fogging (household insecticidal spraying), or individual prevention behaviors occur.

The objective of this study is to identify relationships between household social-ecological factors and *Aedes aegypti* presence and dengue cases, respectively, in Machala, Ecuador. We also test if *Aedes aegypti* presence is an indicator of household dengue fever cases. Household characteristics associated with the dengue fever transmission cycle are potentially identifiable targets for public health vector control intervention to reduce mosquitoes.

## Methods

*Cluster study design* – The data used in this study were collected as part of a SUNY Upstate Medical University epidemiological study of dengue in Machala, Ecuador conducted from 2014–2016. Four sentinel clinics operated by the MSP were selected for passive detection of dengue cases used to enroll households into the study. These clinics were chosen based on historical patterns of dengue detection and available medical resources. A cluster study design was used to enroll participants. Index cases were randomly selected from clinically diagnosed dengue fever cases passively detected in sentinel healthcare facilities. After identifying index households, up to four additional households were enrolled within a 200m radius of each index house, representing the approximate flight range of *Ae. aegypti* mosquitoes; this study protocol has been described in detail previously [38]. Each household was visited once during the three-year study period, at which time household survey data, entomological data, and blood samples from individuals were collected. Locations of enrolled households (i.e. latitude and longitude coordinates) were recorded using Garmin handheld global positioning system (GPS) units.

*Household survey* – Field teams comprising local Ecuadorians visited each enrolled household, administering a survey to the head of household. Survey questions were developed based on previous studies from the region, which identified potential local factors of interest as they relate to risk of dengue transmission [30]. The survey was used to collect information on household demographics, socioeconomic status, access to municipal services, and human behaviors that may influence the presence of mosquitoes (e.g. water storage practices and use of pesticides). Field teams also visually assessed housing conditions and structures, collecting data on patio conditions, window screening, and construction materials. General housing condition was ranked by technicians on site, ranging from “good” housing, which consisted of new construction in good repair (e.g. no holes in walls, maintained roof, etc.), to “poor” conditions where structures were not maintained (e.g. older buildings in disrepair, observable damage to structure, etc.).

*Entomological survey* – Field teams of entomology technicians visited households enrolled in the cluster study from 2014–2016. Households were surveyed for mosquitoes within two days of enrollment to assess abundance and presence of *Ae. aegypti*. Sampling consisted of backpack aspirator (Prokopack) collection of adult mosquitoes in and around households [39]. Adult mosquitoes were stored in coolers in the field, then identified, counted, and sorted by sex and species (i.e. *Ae. aegypti* versus other) at the entomological lab at the Technical University of Machala. Abundance counts of female *Ae. aegypti* for each year of the study were aggregated by households into a binary (i.e. presence/absence) dataset for statistical modeling.

*Human case data* – Data on dengue infection status were collected for enrolled households from 2014–2015; diagnostic results were not available for 2016. The study protocol for collection of human case data was reviewed and approved by the Institutional Review Boards at the State University of New York (SUNY) Upstate Medical University, Cornell University, the Human Research Protection Office of the U.S. Department of Defense, the Luis Vernaza Hospital in Guayaquil, Ecuador, and the Ecuadorian MSP. Prior to initiating the study, all participants, or adult legal guardians, engaged in a written informed consent process. Cases detected via passive clinical surveillance were informed that they may be randomly selected to participate in the cluster study prior to signing the informed consent. The study included children (> 6 months) to adults who were evaluated for DENV infection.

Laboratory diagnostic tests for DENV were run on blood samples collected from study participants. These included NS1 rapid strip tests (Panbio Dengue Early Rapid), ELISA assays for NS1 (Panbio Dengue Early ELISA), IgM immunoassays (Panbio Dengue Capture IgM), and RT-PCR. Full details of the diagnostic testing used in this study are outlined in Stewart-Ibarra et al. (2018) [27]. Household members were considered positive for DENV infection if at least two diagnostic tests were positive. Case counts for each given year of the study were aggregated by households into a binary dataset for statistical modeling.

*Statistical Analysis* - Data from households enrolled in the Machala study were assessed for the possible influence of the clustered study design on data structure, ensuring the selection of an appropriate statistical modeling framework. Intraclass correlation coefficient values indicated that household clusters did not impact observed variance in the data [40]. Consequently, logistic regression with multimodel selection was conducted to identify factors influencing the household presence of *Ae. aegypti* mosquitoes or dengue cases, respectively. We employed a generalized linear model (GLM) statistical framework, specifying a logistic modeling distribution (GLM, family = binomial, link = logit), to identify household-level influences on outcomes of interest (i.e. mosquito presence or presence of dengue cases). Model searches were performed in R (ver. 3.6.2), using the ‘glmulti’ package for multimodel selection, and run until convergence using glmulti’s genetic algorithm (GA) [41]. The top models from each search were ranked via Akaike Information Criterion, corrected for small sample size (AICc).

Variance inflation factors (VIF) were calculated to evaluate multi-collinearity in household survey variables, with values below 10 indicating low collinearity [42]. Stability of top models was assessed via condition numbers (k), where values below 30 indicate stable models [43]. Global Moran’s I with inverse distance weighting was performed in ArcMap (ver. 10.6.1, ESRI, Redlands, CA, USA) on the residuals from the best-fit models to test for presence of spatial autocorrelation, a violation of GLM assumptions [44].

## Results

Ninety-one cases with lab-confirmed acute symptomatic DENV infections were selected for this study via passive surveillance at MSP sentinel facilities during the 2014–2016 study period. These index cases were used to subsequently enroll an additional 369 households into the cluster study, for a total of 460 households in Machala, spanning 91 sampling clusters surveyed for household risk factors and mosquito presence during the study (Fig. 1C, Fig. 2).

**Figure 2.**
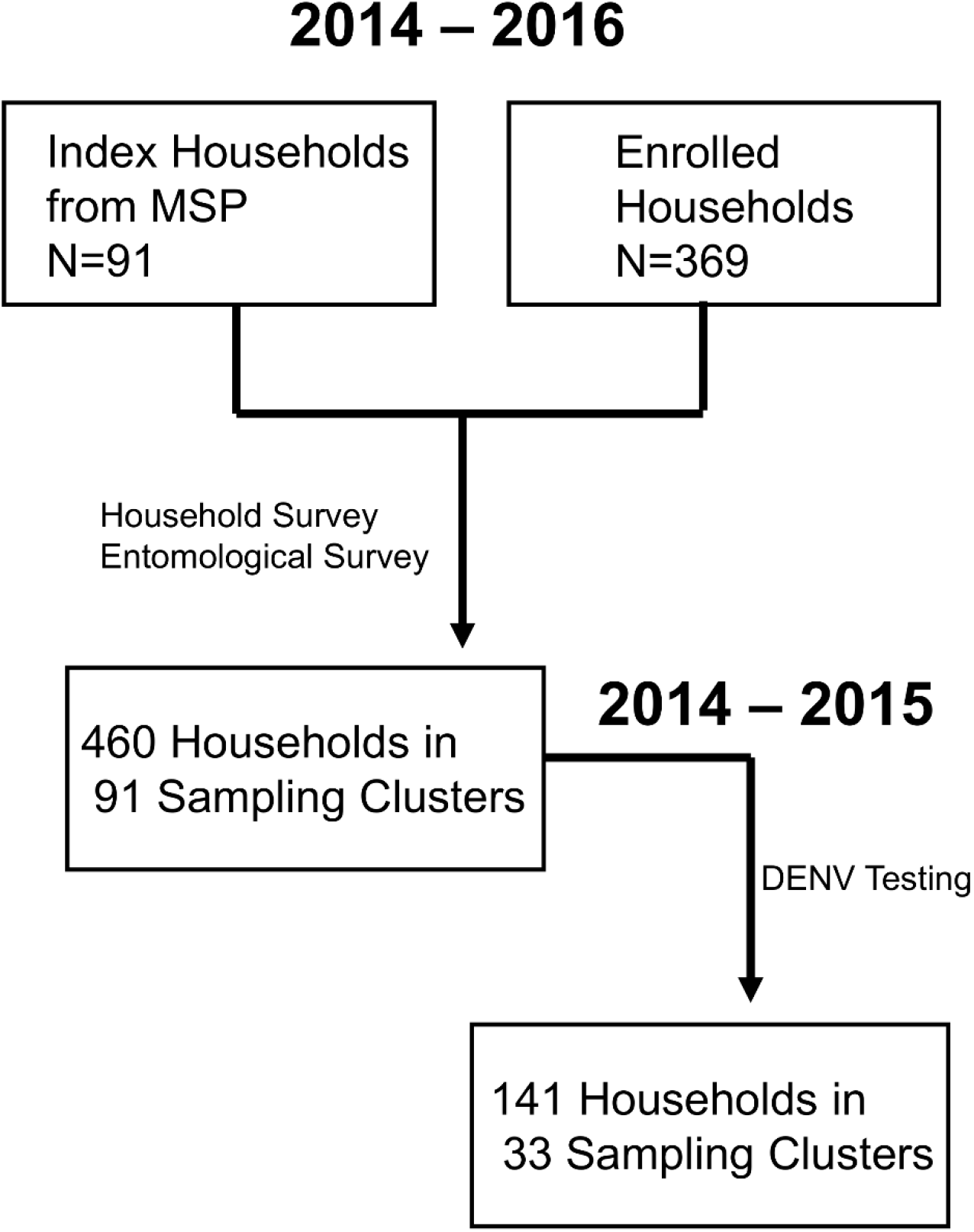
Diagram of household enrollment and data collection for cluster study design in Machala, Ecuador.

*Aedes aegypti* mosquitoes were collected in 54.1% of households (n=249), and 43.9% of households (n=202) had female *Ae. aegypti* present. Summary household surveys results are presented in Table 1.

**Table 1.** Summary of social-ecological variables collected from 2014–2016 household surveys in Machala, Ecuador. Dengue survey households represent a subset of households where entomological surveys were conducted.

| Parameter | Households<br>(n=460) | % | Households<br>(n=141) | % |
| --- | --- | --- | --- | --- |
|  | Entomological Survey |  | Dengue Survey |  |
| Housing conditions |  |  |  |  |
| Good Condition Housing | 144 | 32.0% | 55 | 39.0% |
| Poor Condition Housing | 55 | 12.0% | 16 | 11.3% |
| Cane House Construction | 32 | 7.2% | 9 | 6.4% |
| Wood House Construction | 9 | 2.0% | 2 | 1.4% |
| House is Rented | 40 | 9.5% | 15 | 10.6% |
| Piped Water in Household | 320 | 76.0% | 100 | 70.9% |
| Interruptions in Piped Water | 245 | 53.3% | 78 | 55.3% |
| Municipal Garbage Collection | 400 | 95% | 130 | 92.2% |
| Municipal Sewage | 356 | 84.8% | 119 | 84.4% |
| Septic Tank | 54 | 12.8% | 20 | 14.2% |
| Air Conditioning | 30 | 7.1% | 13 | 9.2% |
| Uses a Fan | 164 | 39.0% | 70 | 49.6% |
| Screens on Windows | 102 | 22.8% | 44 | 31.2% |
| Window Screen in Good Condition | 88 | 19.7% | 35 | 24.8% |
| Access to Paved Roads | 278 | 62.2% | 68 | 48.2% |
| Standing Water Present | 213 | 49.5% | 72 | 51.1% |
| Adjacent to Abandoned Housing | 119 | 26.6% | 49 | 34.8% |
| Patio Present | 367 | 82.1% | 111 | 78.7% |
| Patio in Bad Condition | 81 | 18.1% | 21 | 14.9% |
| Patio Shaded (> 50%) | 56 | 12.5% | 19 | 13.5% |
| Household Demographics and Practices |  |  |  |  |
| Head of Household (HOH) Employed | 359 | 85.7% | 120 | 85.1% |
| HOH male | 323 | 76.7% | 103 | 73.0% |
| HOH Earns Less than Minimum Wage | 74 | 17.6% | 11 | 7.8% |
| HOH Has Secondary Education | 161 | 38.2% | 57 | 40.4% |
| Stores Water | 283 | 67.2% | 96 | 68.1% |
| Know mosquitoes transmit dengue | 372 | 88.8% | 124 | 87.9% |
| Know standing water produces mosquitoes | 402 | 95.9% | 136 | 95.7% |
| Uses Larvicide | 98 | 23.3% | 47 | 33.3% |
| Uses Abate | 151 | 35.9% | 2 | 1.4% |

Housing conditions vary considerably in Machala, with approximately one third (32%) of surveyed housing classified in good condition, and 12% of housing structures classified in poor condition. Access to municipal services among sampled households was high, with 62.2% accessible by paved roads, 76.0% having access to piped water, 84.8% with access to sewage, and 95.0% receiving garbage collection services. The head of household was employed in 85.3% of surveyed households, and 17.6% of heads of household earned less than the minimum wage.

Four hundred study participants from enrolled households were tested for dengue infection status during 2014-2015, and 311 of these had valid diagnostic test results and complete household and entomological survey data. Infection status during 2014–2015 was aggregated to the household level for 141 households in 33 sampling clusters (Fig. 1D). Of these households, 29.08% had one or more household members positive for dengue. All four DENV serotypes were detected in Machala during the study period, and of the thirty-two households tested for dengue serotype, DENV-2 (56.3%) and DENV-1 (40.6%) were the most prevalent infections. *Aedes aegypti* mosquitoes were collected in 57.4% of households (n=81) that were tested for dengue infection, and 49.6% of these households had female *Ae. aegypti* present (n=70). Model selection for household presence of female *Ae. aegypti* in 2014–2016 yielded 53 models within 2 AICc units of the best fit (top ranked) model, indicating a lack of easily distinguishable informative explanatory variables. The number of people in the household, presence of paved roads, and water interruptions were statistically significant risk factors associated with female *Aedes aegypti* presence in the top-ranked model; housing constructed of bamboo cane and houses with air conditioning were found to be protective against mosquito presence (Table 2). Effect sizes in the model were small, and model instability was high (k=360.94).

**Table 2.** Best-fit logistic regression model of household female *Aedes aegypti* presence in Machala, Ecuador.

| Model | Estimate | SE | P-Value | OR | 95% CI |
| --- | --- | --- | --- | --- | --- |
| Intercept | -2.79 | 0.73 | < 0.001 |  |  |
| HoH Age | 0.01 | 0.01 | 0.106 | 1.01 | 0.99 – 1.03 |
| No. People in Household | 0.14 | 0.06 | 0.014 | 1.15 | 1.04 – 1.29 |
| House in Poor Condition | 0.65 | 0.37 | 0.077 | 1.84 | 0.91 – 3.86 |
| Cane Housing | -1.11 | 0.50 | 0.028 | 0.28 | 0.10 – 0.72 |
| Wood Housing | -1.07 | 0.76 | 0.160 | 0.37 | 0.07 – 1.54 |
| Screens in Good Condition | -0.53 | 0.27 | 0.051 | 0.59 | 0.34 – 1.01 |
| Paved Roads | 0.46 | 0.23 | 0.042 | 1.70 | 1.08 – 2.68 |
| Water Interruptions | 0.58 | 0.21 | 0.007 | 1.76 | 1.16 – 2.68 |
| Trash Collection | 1.02 | 0.60 | 0.087 | 0.99 | 0.62 – 1.57 |
| Air Conditioning | -1.39 | 0.52 | 0.007 | 0.25 | 0.08 – 0.63 |

AICc=548.86, pseudo $R^2=0.16$

Model selection for household presence of dengue cases in 2014–2015 yielded 26 models within 2 AICc units of the best fit (top ranked) model. Housing constructed of cane, presence of nearby abandoned houses, shaded patios, HOH employment status, and air conditioning were statistically significant risk factors for dengue cases (Table 3). Households classified in good condition were protective. Presence of female *Ae. aegypti* was not selected as a predictor in the best fit model for household dengue cases, but was included as a variable in 22.2% of models within 2 AICc units of the best model, but never achieved statistical significance. The best fit model’s low condition number (k=5.46) indicated model stability.

**Table 3.** Best-fit logistic regression model of household presence of dengue cases in Machala, Ecuador.

| Model | Estimate | SE | P-Value | OR | 95% CI |
| --- | --- | --- | --- | --- | --- |
| Intercept | -3.21 | 0.98 | 0.001 |  |  |
| House in Good Condition | -1.37 | 0.52 | 0.009 | 0.26 | 0.09 – 0.71 |
| House Made of Cane | 1.55 | 0.85 | 0.068 | 4.69 | 0.89 – 24.65 |
| Adjacent to abandoned property | 1.05 | 0.45 | 0.020 | 2.86 | 1.18 – 6.94 |
| Shaded Patio | 1.75 | 0.61 | 0.004 | 5.77 | 1.75 – 19.04 |
| HOH Works | 2.04 | 0.94 | 0.030 | 7.71 | 1.21 – 48.89 |
| Air Conditioning | 1.51 | 0.69 | 0.028 | 4.53 | 1.17 – 17.51 |

AICc=156.12, pseudo $R^2=0.23$

Intraclass correlation (ICC) values for both entomological and human case datasets were extremely low (i.e. near or equal to zero), indicating that the clustered study design did not meaningfully contribute to variance in the data. Results from the VIF analysis showed that only two variables from the household survey – brick housing construction and primary education –contributed to model collinearity (i.e. VIF ≥ 10), which can lead to unreliable estimates of error for regression coefficients; these variables were excluded from model searches. Despite the clustered arrangement of households in the study design, global clustering was not detected in best fit model residuals for mosquito presence (Moran’s I= 0.06, p-value=0.17) or dengue presence (Moran’s I= −0.13, p-value=0.18), indicating no impact of spatial autocorrelation on model results.

## Discussion

This study provides a household-level assessment of dengue fever risk in Machala, Ecuador, a city with historically high burden of the disease. Indicators of mosquito presence are at times used by public health agencies as a proxy for risk of disease exposure and basis for vector control operations, particularly in absence of active human surveillance data [2,6]. This is an intuitive approach, as widespread surveillance of human cases is resource intensive, and vector presence is a necessary component of transmission cycles for pathogens like dengue virus [45,46]. Identification of factors that promote mosquito presence would provide valuable information for creating targeted interventions. For example, a recent study in Huaquillas, Ecuador, an arid city to the South of Machala on the Peruvian border, found associations between household *Ae. aegypti* presence and the reliability of water services and septic systems, respectively, which could be leveraged into actionable abatement strategies [47]. While *Ae. aegypti* is closely associated with built urban and human environments, we could not identify distinct links between increased risk of mosquito presence and household risk factors assessed here, even with detailed, household-level data across multiple clusters of households spanning a range of social-ecological conditions in the city. This analysis did not yield stable models for mosquito presence, suggesting that measured social risk factors and personal protective behaviors did not reliably indicate local presence of female *Ae. aegypti* in Machala. In absence of identifiable household targets for intervention, following current MSP advisories and educational initiatives for larval source reduction of container-breeding mosquitoes is recommended [2,26].

Unlike mosquito presence, dengue infection status of households in Machala was connected to variables describing housing conditions. Household factors that influenced presence of dengue fever were broadly consistent with other studies from this region, where housing conditions across multiple scales have been linked to transmission risk in Ecuador [14,20,30,34,43,48]. Housing structure in good condition was identified as the top protective factor against dengue presence, a finding which has been observed in other studies conducted in coastal Ecuador [20,30]. Houses in good repair limit exposure to mosquitoes, and the association between substandard housing structure and arboviral risk has been repeatedly documented in locations beyond Ecuador [49-51]. Houses made of cane were a risk factor for dengue, which is consistent with typical housing found in low income areas with active dengue transmission in Ecuador [52]. Generally found in the urban periphery of Machala, housing constructed from cane does not exclude potentially infectious mosquitoes from entering households [12]. Adjacency to abandoned properties and presence of shaded patios were identified as risk factors in this study, and have also been linked to transmission risk in other studies in the region, promoting favorable habitat and oviposition sites for container-breeding mosquitoes [14,48].

Air conditioning and HOH employment were risk factors for dengue presence in this study. These relationships are counterintuitive, as these factors are typically regarded as protective against exposure to mosquitoes in other systems [53–55]. Air conditioning eliminates the need to open door and windows for ventilation, reducing exposure to mosquitoes. Head of household employment is often a marker for higher socioeconomic status, which is generally associated with improved housing quality and lower risk [6,51]. Although the survey tool used in this study represents a major step forward in the collection of fine-scale data regarding risk of dengue transmission in Ecuador, the questions did not necessarily capture the nuance of some risk elements. For example, MSP technicians in Machala have observed that families will only run air conditioning units at night, relying on open windows to cool houses during the day (*pers. comm.)*. Patterns of human behavior not captured in the survey could account for the counterintuitive relationship; similarly, the household survey tool did not account for events outside of the home, where exposures associated with travel and employment may occur. Such an association was reported in the Galapagos, where travel patterns were associated with self-reported dengue illness [43].

A previous study, conducted by Kenneson et al. (2017), also examined household dengue fever risk in Machala during 2014–2015 [48]. In the current study, we restricted analyses to households with complementary dengue case and entomological survey data, and used a more conservative dengue case definition, requiring at least two positive diagnostic tests, excluding IgG immunoassay results, which indicate longer term exposures [4]. The 2017 study, with broader case definitions and a larger group of households, also found that adjacency to abandoned properties and shaded patios were significant risk factors for dengue fever, but also identified frequent water interruptions as a significant risk factor. We found that frequent water interruptions was selected as a risk factor in the best-fit (but unstable) model for *Ae. aegypti* presence, but was not selected for in the best-fit model for dengue case presence, using the more conservative case definition. The 2017 study did not examine overall housing condition, making direct comparisons of findings unfeasible, but instead demonstrated that preventive actions and perceptions were negatively associated with dengue fever outcomes. While these studies leverage overlapping sampling events, the differences in objectives, case definitions, and factors analyzed, indicate a need for further examination of the implications for connections in the system between the transmission conditions, the scale of human behavior and household impact, and which components of the lived environment interact with the vector-human transmission cycle.

Interestingly, female *Aedes aegypti* presence was not identified as a significant risk factor for household dengue infection status in the top ranked models in this study. Mosquito abundance data have been shown in some instances to correlate with risk of dengue infection, particularly in longitudinal studies that can account for temporal variability and lagged effects [18,56]. Yet, indoor household *Ae. aegypti* densities can be notoriously low, and samples collected with backpack aspirators are often highly dependent on factors such as user skill and complexity of the indoor environment [57-59]. Furthermore, some studies suggest that alternative trapping methods, such as the use of passive BG-Sentinel traps, may be better suited for targeting *Ae. aegypti* [9,58,59]. In this study, we did not use mosquito abundance as a dependent variable in modeling due to low counts of adult mosquitoes (e.g. <10) recorded in household surveys. Nevertheless, backpack aspiration is a logistically feasible surveillance method for many vector control agencies in Latin America, and there is evidence of high sensitivity for detecting the presence of mosquitoes in households [18,39,60]. Also notable is the lack of association between household dengue cases in this study and indicators of water availability. Frequent water interruptions promote water storage behaviors, thus impacting mosquito reproduction and abundance, and interruptions in water service was selected as a predictor in the unstable best-fit model for mosquito presence in this study [61]. Access to piped water and water storage have been linked to both dengue infections and the proliferation of *Ae. aegypti* across multiple spatial scales, in Machala and other coastal Ecuadorian cities [14,20,30,31,47,62]. The lack of association between mosquito presence and dengue infections indicated in this study may instead reflect a disconnect between household conditions and locations where transmission is occurring, particularly in the context of hyperendemic dengue transmission in Machala. Studies conducted in other Latin American cities with hyperendemic dengue transmission have shown that within cities, transmission risk is influenced by neighboring census tracts, and no specific locations within the city were driving dengue cases [63,64]. Daily human movements and social connections within hyperendemic spaces can also drive dengue transmission, particularly on fine spatial scales [65,66]. Given the high prevalence of the disease in the community, risk of transmission in Machala may be more dependent on social and behavioral factors that increase exposure to infective mosquito bites, rather than mere presence or absence of the vector in the immediate vicinity of the household. Although not informative at the household level in this study, municipal and sub-regional entomological surveillance is still a cornerstone of successful public health vectors control programs, where mosquito counts are necessary to set goals and proactively trigger the deployment of vector control services ahead of epidemic peaks [6,67,68]. In locations with hyperendemic arboviral transmission, like Machala, surveillance of human cases may provide better primary information for guiding policy and developing community interventions on fine spatial scales.

Household-level data are useful in advancing our understanding of localized disease transmission and exposure risk, yet there are caveats that must be considered before findings are used in an applied capacity. The statistical models used in this study are correlative, and we therefore must avoid assigning causality to any significant relationships without further investigation. Furthermore, data collected via household surveys certainly provide information on a fine scale, but they do not capture the full social-ecological transmission environment. This is exemplified by the lack of information concerning potential workplace and school exposures, which are part of the greater transmission social-ecological system at this scale. We also caution against reducing localized risk indicators into a dichotomy of “poor” versus “affluent” neighborhoods. This reductionist approach does not account for nuances in risk due to personal behaviors, or exposure due to hyperendemic presence of mosquito vectors. Focusing solely on economic indicators of risk has the additional potential to generate stigma in communities, which can undermine public health initiatives [69].

The results of this study aid us in identifying areas for further study as potential targets for developing proactive mosquito control activities and intervention strategies in Machala. The indication of missing linkages between entomological exposure risk indicators at the household level, where vector control intervention is often carried out, and the indicators of dengue infection, point to a need for a more complete examination of the social-ecological transmission environment. Nonetheless, comparing our findings to previously published work, we have identified commonalities in risk factors across spatial scales. Thus, we indicate that some household-level risk factors, such as condition of housing structure, may be generalizable in a policy development capacity.

## Data Availability

Since the publication includes confidential patient information, we cannot make the datasets fully available with the paper. This would be in violation of confidentiality as expressed in the Informed Consent approved by U.S. and Ecuadorian IRBs. Data are thus available from the Ethics Committee of the SUNY Upstate Medical University for researchers who meet the criteria for access to confidential data. The de-identified datasets in the current study are available from Lisa Ware at on reasonable request.

## Acknowledgments

Many thanks to the SUNY Upstate field technicians and other staff at the Ministry of Health of Ecuador, who generated the data used in this analysis. We thank our local field team and coordinators in Machala for their dedication and support: Jefferson Adrian, Victor Arteaga, Jose Cueva, Reagan Deming, Carlos Enriquez, Prissila Fernandez, Froilan Heras, Naveed Heydari, Jesse Krisher, Lyndsay Krisher, Elizabeth McMahon, Eunice Ordoñez, Tania Ordoñez, and Rachel Sippy.

## Notes

### Competing Interest Statement

The authors have declared no competing interest.

### Clinical Trial

Not a clinical trial

### Funding Statement

The author(s) received no specific funding for this work.

### Author Declarations

The study protocol for collection of human case data was reviewed and approved by the Institutional Review Boards at the State University of New York (SUNY) Upstate Medical University, Cornell University, the Human Research Protection Office of the U.S. Department of Defense, the Luis Vernaza Hospital in Guayaquil, Ecuador, and the Ecuadorian MSP.

